# Neonatal deep medullary venous thrombosis radiographic severity is associated with neurodevelopmental impairment

**DOI:** 10.1101/2025.02.07.25321901

**Authors:** Ria Pal, Gabrielle Russo Barsh, Ingrid Luo, Hisham Dahmoush, Sarah Lee, Elizabeth Mayne

## Abstract

Deep medullary vein thrombosis (DMVT) is an increasingly recognized etiology of neonatal brain injury, but remains poorly understood. Our study aimed to assess the association between MRI severity and neurodevelopmental impairment (NDI) in neonates with DMVT, and develop a novel MRI grading system that might inform clinical outcomes. We retrospectively reviewed relevant charts from infants admitted to our tertiary care hospital between January 1990 to March 2023, and evaluated clinical characteristics, MRI features, and neurodevelopmental assessments of this cohort. We developed and validated a simple MRI grading system based on injury severity, categorizing lesions into mild, moderate, or severe groups. Of the 63 neonates with a diagnosis of DMVT, 41 had moderate or severe MRI lesions; those patients were 24-fold more likely to experience NDI compared to those with mild injury (adjusted OR 24.3, 95% CI 4.7-180.2, p<0.001). Of the 52 infants with follow-up data, 40.4% developed NDI; MRI severity was the strongest predictor of impaired outcomes, independent of clinical factors including gestational age, Apgar score and seizures at presentation. Our findings suggest that this pragmatic MRI grading scheme may offer clinicians and researchers a valuable classification and prognostication tool.

## Introduction

The neonatal period is a time of high risk for neurovascular injury.^1,2^ This can include either ischemic injury or venous strokes, which account for 7-30 percent of neonatal strokes. Although features of neonatal cerebral sinus venous thrombosis (CSVT) have been well-described, advancements in MRI technology have led to increasing awareness of thrombosis or congestion of the deep medullary veins.^3,4^ The deep medullary veins drain the central white matter, converging into larger collector vessels that ultimately empty into the subependymal veins of the lateral ventricles.^5^ Deep Medullary Vein Thrombosis (DMVT) can occur with or without CSVT, and has been specifically associated with neonates, though scant existing literature makes it difficult to estimate the true incidence, particularly of isolated DMVT. Descriptions of DMVT have been primarily limited to case reports and small single center case series, which have suggested an association with neurodevelopmental impairment.^5–7^

Radiographically, DMVT is best visualized with MRI, and is characterized by engorgement of the DMVs on T2-weighted or SWI sequences.^8–10^ DMVT can lead to parenchymal injury, but there is a lack of consensus on common patterns of injury and radiological risk factors for neurodevelopmental impairment.^11^

The aim of our study was to analyze demographic, clinical, and radiographic features of our large single-center neonatal DMVT cohort and to create a simple MRI grading system incorporating multiple patterns of brain injury that might inform neurodevelopmental outcomes in this population.

## Methods

This is a retrospective, single-center cohort study examining the association between imaging results and neurodevelopmental outcomes in patients with DMVT.

### Institutional Review Board (IRB) Approval

This study was approved by the Institutional Review Board at Stanford University (IRB number: IRB-69590), with approval granted on 04/04/23. The study adhered to ethical guidelines concerning patient data privacy and confidentiality. As a retrospective, minimal-risk study, this research was exempt from requiring informed consent.

### Patient Selection

We searched the Stanford Sectra PACS, containing all radiology reports from Lucile Packard Children’s Hospital (LPCH) and Stanford Health Care (SHC), for cases of deep medullary venous thrombosis or congestion from January 1990 to March 2023 using the terms “deep medullary venous thrombosis” and “deep medullary vein congestion.” We searched the Stanford imaging database for cases of deep medullary venous thrombosis or congestion from January 1990 to March 2023 using the terms “deep medullary venous thrombosis” and “deep medullary vein congestion.”

Pediatric patients (<18 years) were included if they had MRI available in our imaging database demonstrating deep medullary vein thrombosis or congestion on review by a board-certified pediatric neuroradiologist (H.D.).

### Data Extraction

Patient charts and MRI images were retrieved from the LPCH Epic electronic health record system. Charts were reviewed for pregnancy and delivery history, clinical presentation, MRI findings, treatment, and neurodevelopmental outcomes. Study data were collected and managed using REDCap electronic data capture tools hosted at Stanford University.

### Radiologic Grading

Patients were scanned over this time period on both 1.5T and 3T MRI scanners. Sequences analyzed included T2-weighted images, diffusion weighted images (DWI) with apparent diffusion coefficient (ADC) maps, susceptibility weighted imaging (SWI), and gradient-recalled echo (GRE).

Prior to imaging analysis, a grading scheme was developed by the authors to classify deep medullary venous injury with an emphasis on practical clinical application (Table 1).

**Table 1:**
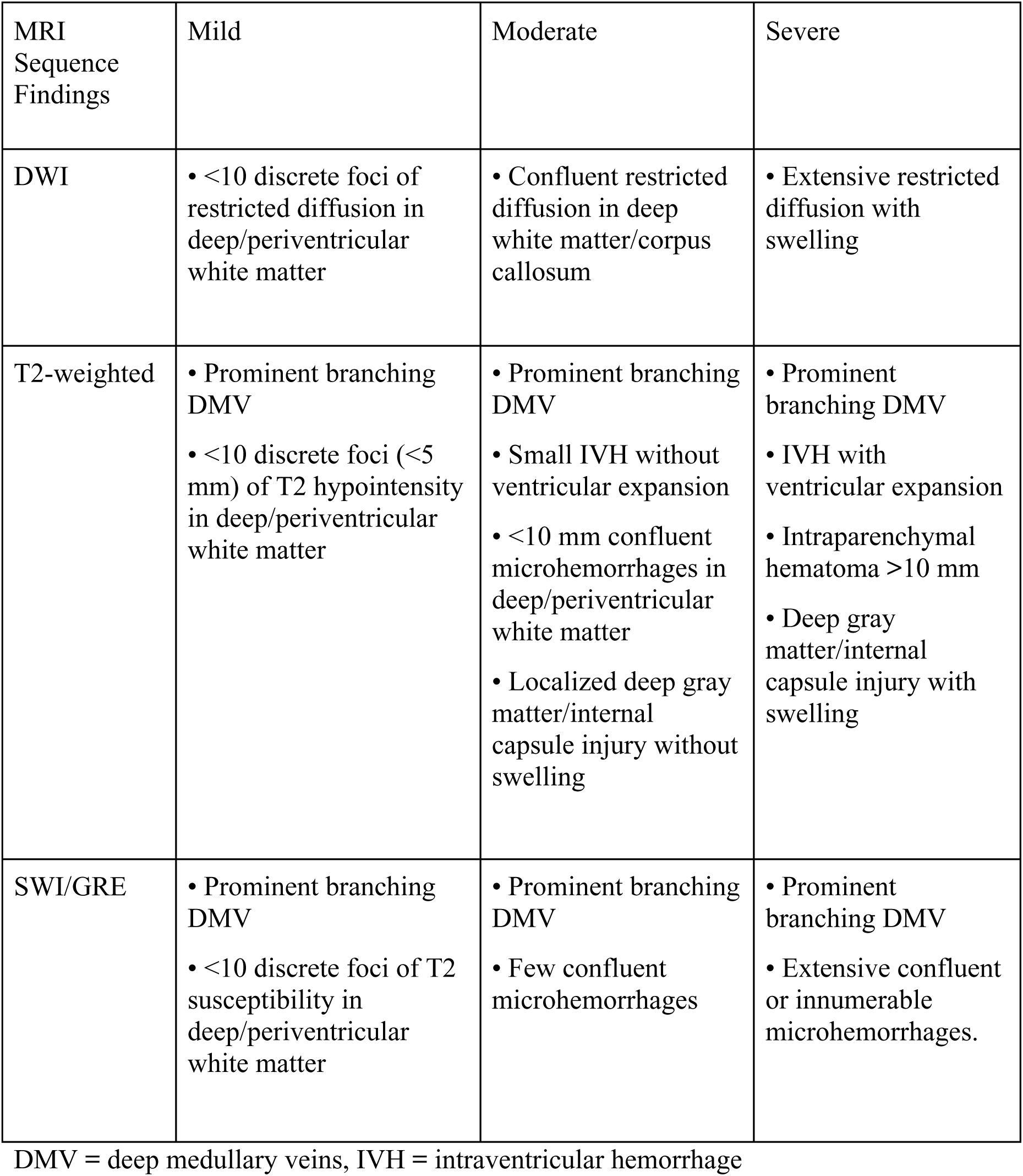
Radiographic Grading System for Deep Medullary Venous Thrombosis.

Initial grading was carried out by two independent graders (R.P. and G.B., board certified child neurologists). Discrepancies between the initial graders were reviewed and resolved with the input of a third grader (H.D.), a board-certified pediatric neuroradiologist. The final grade for each case was determined through consensus among the entire grading team.

### Evaluation of neurodevelopmental impairment

Moderate-severe neurodevelopmental impairment was defined as a composite outcome as previously described after neonatal brain injury.^12^ Survivors were considered to have moderate to severe NDI if they had any of the following: a Bayley score in the language or cognitive domain greater than 2 standard deviations below the mean (specifically, below 70 for the Bayley-II Mental Development Index Score or below 85 for the Bayley III Language or Cognitive Composite Score), a Gross Motor Function Classification System (GMFCS) grade of 2 or higher, severe visual impairment (acuity worse than 20/200 in both eyes, or requiring corrective lenses or surgery for strabismus or blindness), or profound hearing loss (sensorineural hearing loss greater than 30 dB bilaterally, or requiring bilateral auditory amplification). When assessments were obtained serially, the assessment closest to 24 months was analyzed.

### Statistical Analysis

Patient demographics, clinical characteristics, and radiologic findings were stratified based on the MRI severity grades at the time of diagnosis. Continuous variables were reported as median with interquartile range (IQR: 25th percentile, 75th percentile), while categorical variables were expressed as counts and percentages. Between-group differences were evaluated using Kruskal-Wallis test for continuous variables and Fisher’s exact test for categorical variables.

To investigate the association between deep medullary venous thrombosis or congestion and MRI severity at the time of diagnosis, both univariate and multivariate logistic regression analyses were conducted. Robust standard errors were employed to account for potential heteroscedasticity and to provide more conservative estimates of statistical significance. The multivariate model adjusted for hypoxic-ischemic encephalopathy (HIE), congenital heart disease (CHD), gestational age (in weeks), 1-minute Apgar score (dichotomized as <3 or ≥3), seizures at presentation, and mode of delivery (Cesarean or vaginal).

MRI findings were initially scored on a three-tier scale (mild, moderate, and severe) to capture the full spectrum of radiographic findings and enable descriptive analysis for exploratory analyses, such as descriptions of NDI-impaired domains and interval changes between MRI grades. For tests of survival analysis, moderate and severe categories were combined to create a dichotomous variable (mild vs. moderate-severe) to optimize statistical power while maintaining clinical distinctions. This approach was chosen based on preliminary analyses suggesting similar outcomes in moderate and severe groups, and to ensure adequate sample sizes for robust statistical comparison.

To assess the time-dependent association between the development of DMVT and MRI severity at the baseline, survival analysis was conducted using both unadjusted and adjusted approaches. All patients were right-censored at 3 years after diagnosis. Patient-years were measured from the time of initial clinical presentation to either the development of NDI, the last clinical visit, or 3 years post-presentation, whichever occurred first. The cumulative incidence of DMVT was visualized using Kaplan-Meier curves, and adjusted analyses were conducted using a Cox proportional hazards model. The Cox model included the same covariates as the multivariate logistic regression analysis. The proportional hazards assumption was tested using global proportional hazards test. All analyses were performed in R version 4.2.3.

## Results

### Demographics and Clinical Characteristics

Among 64 patients with confirmed deep medullary venous thrombosis or congestion noted on radiology report, one was excluded due to unavailable imaging (performed at outside institution). Baseline characteristics and outcomes for these groups were compared and are summarized in Table 2. Median gestational age was similar between the groups (37.0 weeks, IQR 34.3-38.0 for mild MRI and 37.0 weeks, IQR 34.0-39.0 for moderate/severe MRI). Patients presented at median age two days of life. There were no significant differences in proportions of sex or racial/ethnic distributions (p = 0.228). A higher proportion of patients from the moderate/severe MRI group (56.1%) initially presented with seizures compared to the mild MRI group (27.3%, p = 0.036). Similar rates of hypotonia, encephalopathy, and respiratory distress were observed.

**Table 2:**
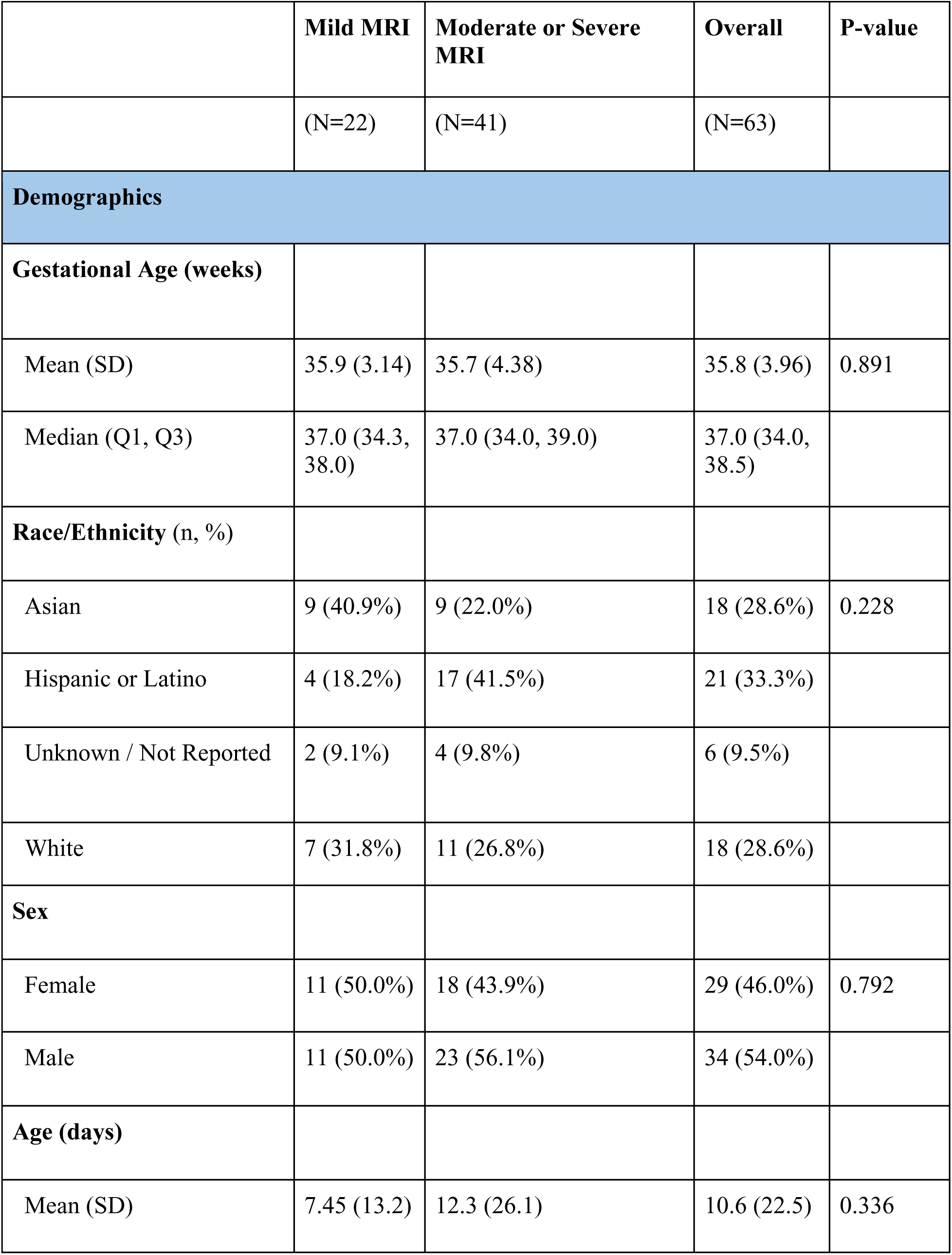
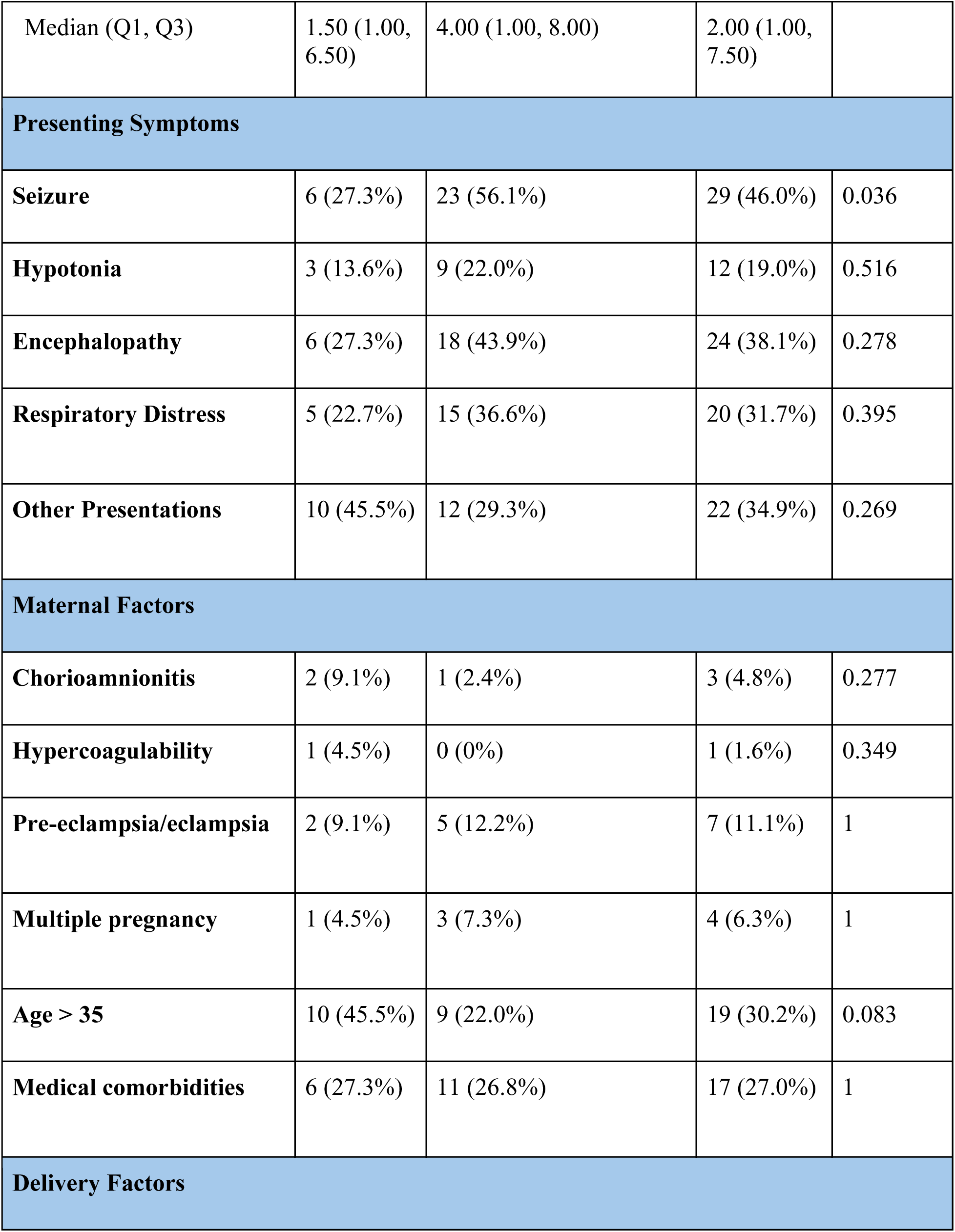
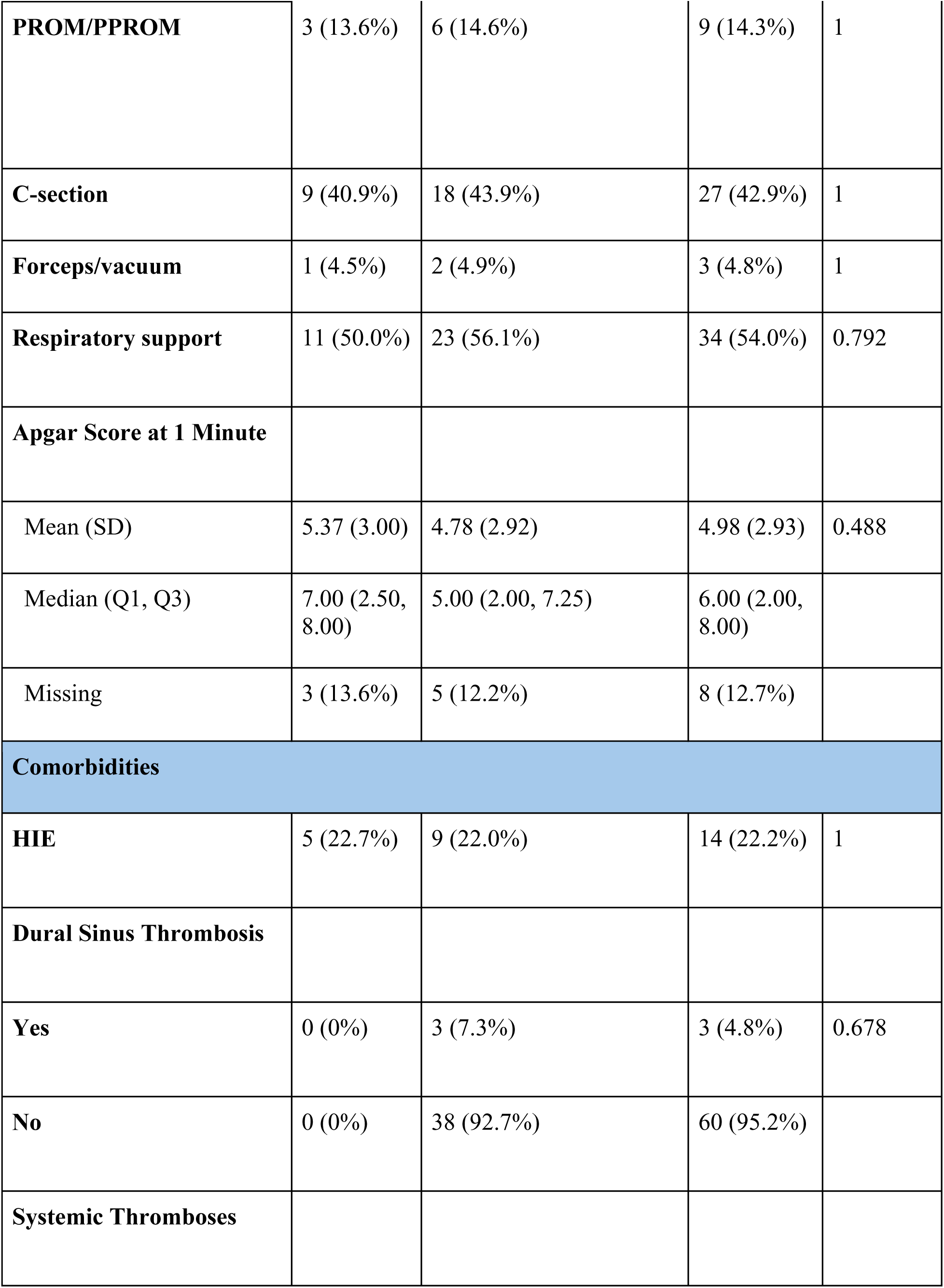
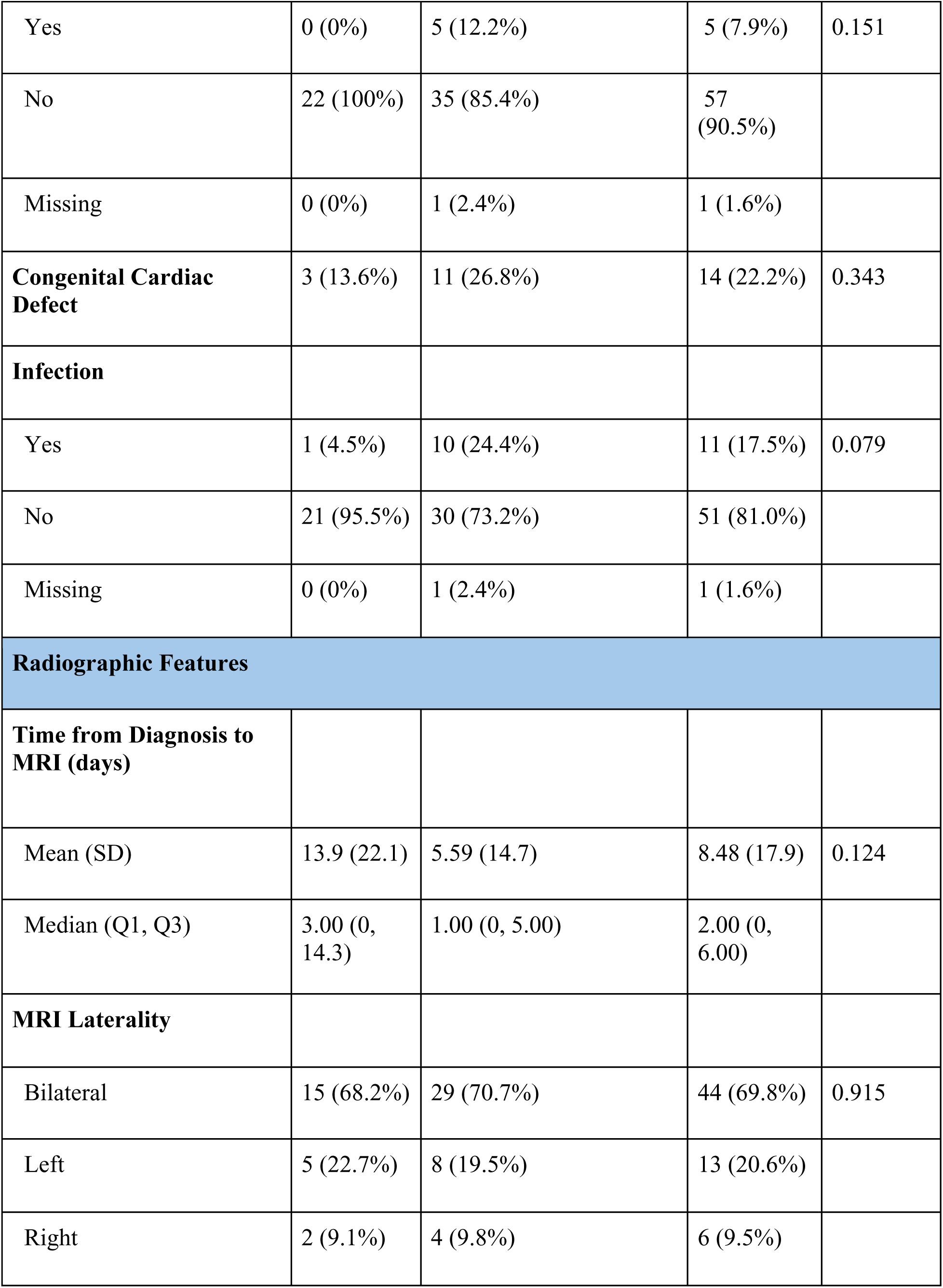
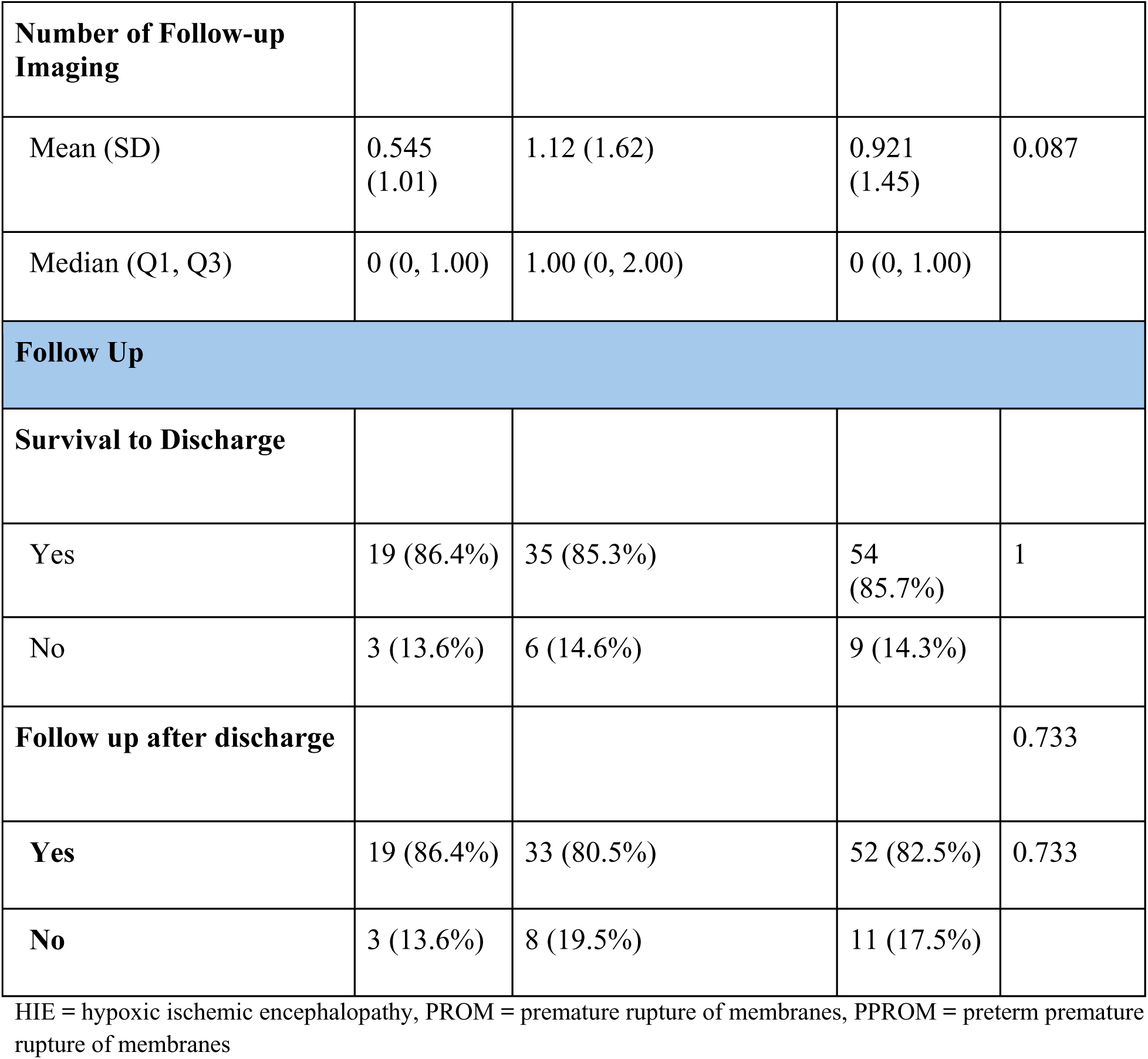
Baseline characteristics and outcomes.

|  | **Mild MRI** | **Moderate or Severe MRI** | **Overall** | **P-value** |
| --- | --- | --- | --- | --- |
|  | (N=22) | (N=41) | (N=63) |  |
| **Demographics** |  |  |  |  |
| **Gestational Age (weeks)** |  |  |  |  |
| Mean (SD) | 35.9 (3.14) | 35.7 (4.38) | 35.8 (3.96) | 0.891 |
| Median (Q1, Q3) | 37.0 (34.3, 38.0) | 37.0 (34.0, 39.0) | 37.0 (34.0, 38.5) |  |
| **Race/Ethnicity (n, %)** |  |  |  |  |
| Asian | 9 (40.9%) | 9 (22.0%) | 18 (28.6%) | 0.228 |
| Hispanic or Latino | 4 (18.2%) | 17 (41.5%) | 21 (33.3%) |  |
| Unknown / Not Reported | 2 (9.1%) | 4 (9.8%) | 6 (9.5%) |  |
| White | 7 (31.8%) | 11 (26.8%) | 18 (28.6%) |  |
| **Sex** |  |  |  |  |
| Female | 11 (50.0%) | 18 (43.9%) | 29 (46.0%) | 0.792 |
| Male | 11 (50.0%) | 23 (56.1%) | 34 (54.0%) |  |
| **Age (days)** |  |  |  |  |
| Mean (SD) | 7.45 (13.2) | 12.3 (26.1) | 10.6 (22.5) | 0.336 |
| Median (Q1, Q3) | 1.50 (1.00, 6.50) | 4.00 (1.00, 8.00) | 2.00 (1.00, 7.50) |  |
| **Presenting Symptoms** |  |  |  |  |
| **Seizure** | 6 (27.3%) | 23 (56.1%) | 29 (46.0%) | 0.036 |
| **Hypotonia** | 3 (13.6%) | 9 (22.0%) | 12 (19.0%) | 0.516 |
| **Encephalopathy** | 6 (27.3%) | 18 (43.9%) | 24 (38.1%) | 0.278 |
| **Respiratory Distress** | 5 (22.7%) | 15 (36.6%) | 20 (31.7%) | 0.395 |
| **Other Presentations** | 10 (45.5%) | 12 (29.3%) | 22 (34.9%) | 0.269 |
| **Maternal Factors** |  |  |  |  |
| **Chorioamnionitis** | 2 (9.1%) | 1 (2.4%) | 3 (4.8%) | 0.277 |
| **Hypercoagulability** | 1 (4.5%) | 0 (0%) | 1 (1.6%) | 0.349 |
| **Pre-eclampsia/eclampsia** | 2 (9.1%) | 5 (12.2%) | 7 (11.1%) | 1 |
| **Multiple pregnancy** | 1 (4.5%) | 3 (7.3%) | 4 (6.3%) | 1 |
| **Age > 35** | 10 (45.5%) | 9 (22.0%) | 19 (30.2%) | 0.083 |
| **Medical comorbidities** | 6 (27.3%) | 11 (26.8%) | 17 (27.0%) | 1 |
| **Delivery Factors** |  |  |  |  |
| **PROM/PPROM** | 3 (13.6%) | 6 (14.6%) | 9 (14.3%) | 1 |
| **C-section** | 9 (40.9%) | 18 (43.9%) | 27 (42.9%) | 1 |
| **Forceps/vacuum** | 1 (4.5%) | 2 (4.9%) | 3 (4.8%) | 1 |
| **Respiratory support** | 11 (50.0%) | 23 (56.1%) | 34 (54.0%) | 0.792 |
| **Apgar Score at 1 Minute** |  |  |  |  |
| Mean (SD) | 5.37 (3.00) | 4.78 (2.92) | 4.98 (2.93) | 0.488 |
| Median (Q1, Q3) | 7.00 (2.50, 8.00) | 5.00 (2.00, 7.25) | 6.00 (2.00, 8.00) |  |
| Missing | 3 (13.6%) | 5 (12.2%) | 8 (12.7%) |  |
| **Comorbidities** |  |  |  |  |
| **HIE** | 5 (22.7%) | 9 (22.0%) | 14 (22.2%) | 1 |
| **Dural Sinus Thrombosis** |  |  |  |  |
| **Yes** | 0 (0%) | 3 (7.3%) | 3 (4.8%) | 0.678 |
| **No** | 0 (0%) | 38 (92.7%) | 60 (95.2%) |  |
| **Systemic Thromboses** |  |  |  |  |
| Yes | 0 (0%) | 5 (12.2%) | 5 (7.9%) | 0.151 |
| No | 22 (100%) | 35 (85.4%) | 57 (90.5%) |  |
| Missing | 0 (0%) | 1 (2.4%) | 1 (1.6%) |  |
| **Congenital Cardiac Defect** | 3 (13.6%) | 11 (26.8%) | 14 (22.2%) | 0.343 |
| **Infection** |  |  |  |  |
| Yes | 1 (4.5%) | 10 (24.4%) | 11 (17.5%) | 0.079 |
| No | 21 (95.5%) | 30 (73.2%) | 51 (81.0%) |  |
| Missing | 0 (0%) | 1 (2.4%) | 1 (1.6%) |  |
| **Radiographic Features** |  |  |  |  |
| **Time from Diagnosis to MRI (days)** |  |  |  |  |
| Mean (SD) | 13.9 (22.1) | 5.59 (14.7) | 8.48 (17.9) | 0.124 |
| Median (Q1, Q3) | 3.00 (0, 14.3) | 1.00 (0, 5.00) | 2.00 (0, 6.00) |  |
| **MRI Laterality** |  |  |  |  |
| Bilateral | 15 (68.2%) | 29 (70.7%) | 44 (69.8%) | 0.915 |
| Left | 5 (22.7%) | 8 (19.5%) | 13 (20.6%) |  |
| Right | 2 (9.1%) | 4 (9.8%) | 6 (9.5%) |  |
| **Number of Follow-up Imaging** |  |  |  |  |
| Mean (SD) | 0.545 (1.01) | 1.12 (1.62) | 0.921 (1.45) | 0.087 |
| Median (Q1, Q3) | 0 (0, 1.00) | 1.00 (0, 2.00) | 0 (0, 1.00) |  |
| **Follow Up** |  |  |  |  |
| **Survival to Discharge** |  |  |  |  |
| Yes | 19 (86.4%) | 35 (85.3%) | 54 (85.7%) | 1 |
| No | 3 (13.6%) | 6 (14.6%) | 9 (14.3%) |  |
| **Follow up after discharge** |  |  |  | 0.733 |
| **Yes** | 19 (86.4%) | 33 (80.5%) | 52 (82.5%) | 0.733 |
| **No** | 3 (13.6%) | 8 (19.5%) | 11 (17.5%) |  |

HIE = hypoxic ischemic encephalopathy, PROM = premature rupture of membranes, PPROM = preterm premature rupture of membranes

### Neuroimaging features

Twenty-two (34.9%) were found to have mild MRI changes, 21 (33.3%) with moderate changes, and 20 (31.7%) with severe changes. The inter-rater reliability for this grading process yielded an 80% concordance rate. Representative images are shown in Figure 1. Time from presentation to MRI was longer in the mild MRI group (median 3 days, mean 13.9 days) compared to the moderate/severe MRI group (median 1 day, mean 5.59 days), though this difference was not statistically significant (p = 0.12). Follow-up imaging studies were more frequent in the moderate/severe MRI group (mean 1.1 studies) compared to the mild MRI group (mean 0.55 studies, p = 0.09).

**Figure 1:**
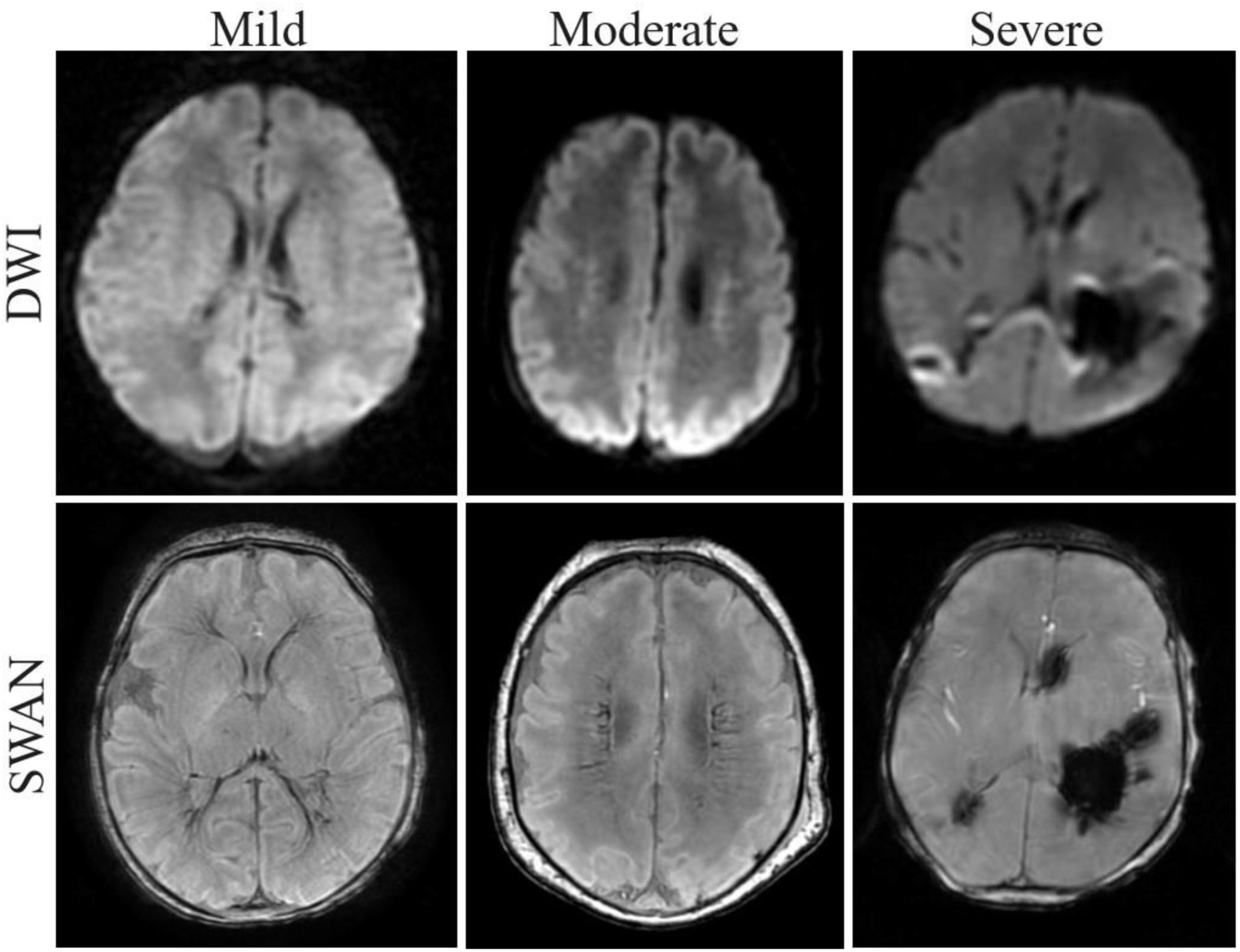
Illustrations of Mild, Moderate, and Severe Classifications in Axial DWI and SWAN Sequences.

### Management and Survival

Nineteen percent (n=12) received hyperhydration for DMVT while 10.9% (n=7) received anticoagulation. All patients who received anticoagulation received heparin (one transitioning to enoxaparin). One was started on heparin for a PICC-associated thrombus, then transitioned to bivalirudin following a cardiac repair. Of the seven who received anticoagulation, all had another indication for anticoagulation, such as systemic thromboses or cerebral sinus venous thrombosis. Survival to discharge rates were similar (86.4% for mild MRI and 85.7% for moderate/severe MRI, p = 1.0). Follow-up visit attendance was also comparable (86.4% in the mild MRI group vs. 80.5% in the moderate/severe MRI group, p = 0.73).

### Neurodevelopmental Outcomes

Of the study cohort, 52 patients (83% of the study cohort) survived to discharge and attended at least one follow up visit with a neurodevelopmental assessment. NDI was found in 21 patients (40.4%) (Table 3). NDI assessment ages varied by evaluation type: cerebral palsy assessments were conducted at median 20 months (IQR: 12-25 months, n=49); hearing assessments at 25 months (IQR: 13-32 months, n=29); vision assessments at 24 months (IQR: 14-43 months, n=24); and Bayley assessments at 22 months (IQR: 14-25 months, n=27). Among the 21 patients with NDI, 2 (9.5%) had mild MRI findings at baseline, each with impairment in a single domain. Seven patients with NDI had moderate MRI findings: two with one impaired domain, three with two impaired domains, and two with three impaired domains. Twelve with NDI had severe MRI findings: eight with one impaired domain, three with two impaired domains, and one with three impaired domains.

**Table 3:**
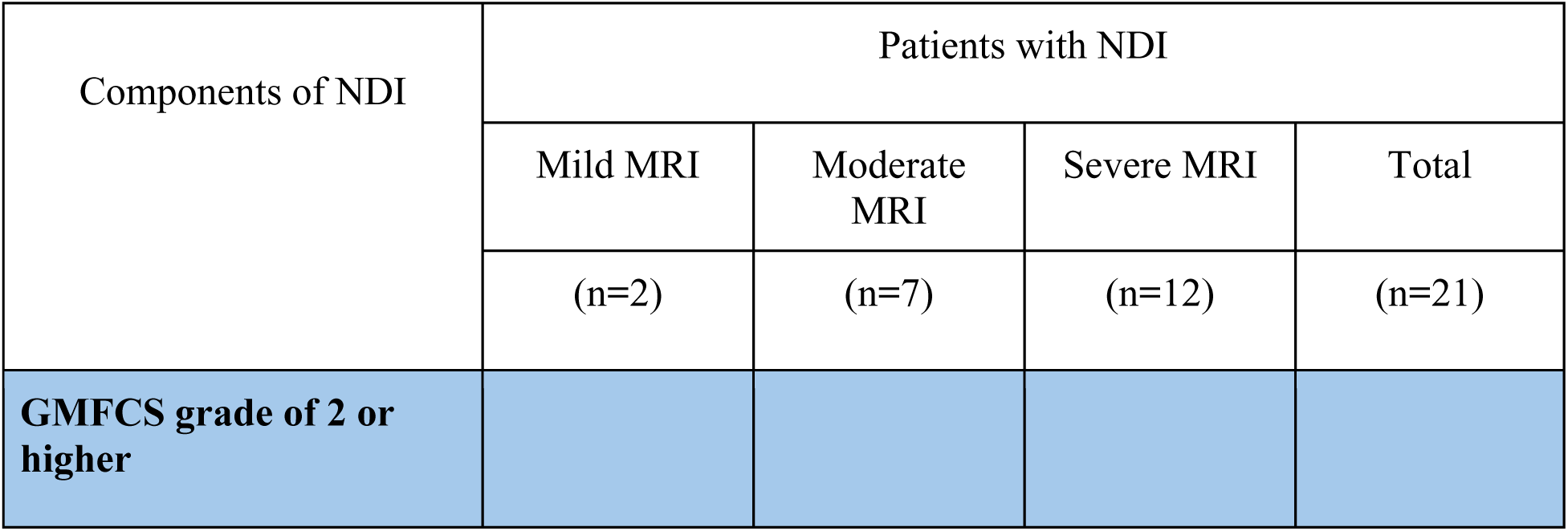
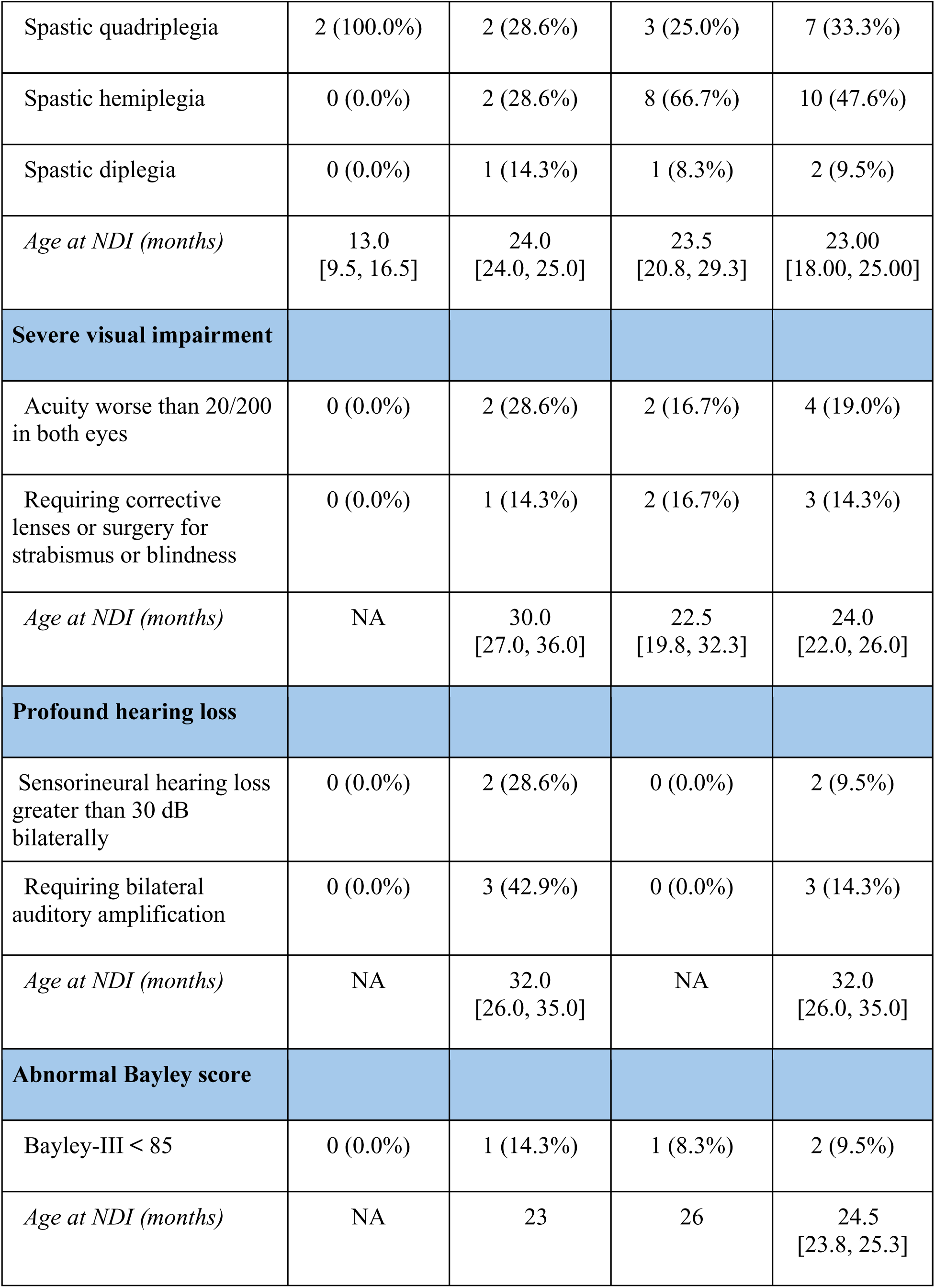
Detailed Evaluation of NDI.

As described above, for the primary survival analysis, we combined the moderate and severe MRI categories into a single “moderate-severe” group. On multivariate linear regression, the odds of NDI were 24.3 times higher in patients with moderate-severe baseline MRI after adjusting for clinical factors (95% CI 4.7-180.2, p<0.001) compared to patients with mild baseline MRI severity, consistent with univariate analysis (OR 12.4, p<0.001). This dichotomization increased statistical power from 71.1% to 81.7% with our sample size of 52 (Figure 2). Patients with mild MRI had a lower probability of developing NDI over 83.1 patient-years (log-rank p=0.040, Figure 3). The median time to NDI development in the moderate-severe group was 2.1 years, while more than 50% of patients with mild MRI severity remained NDI- free throughout the follow-up period.

**Figure 2:**
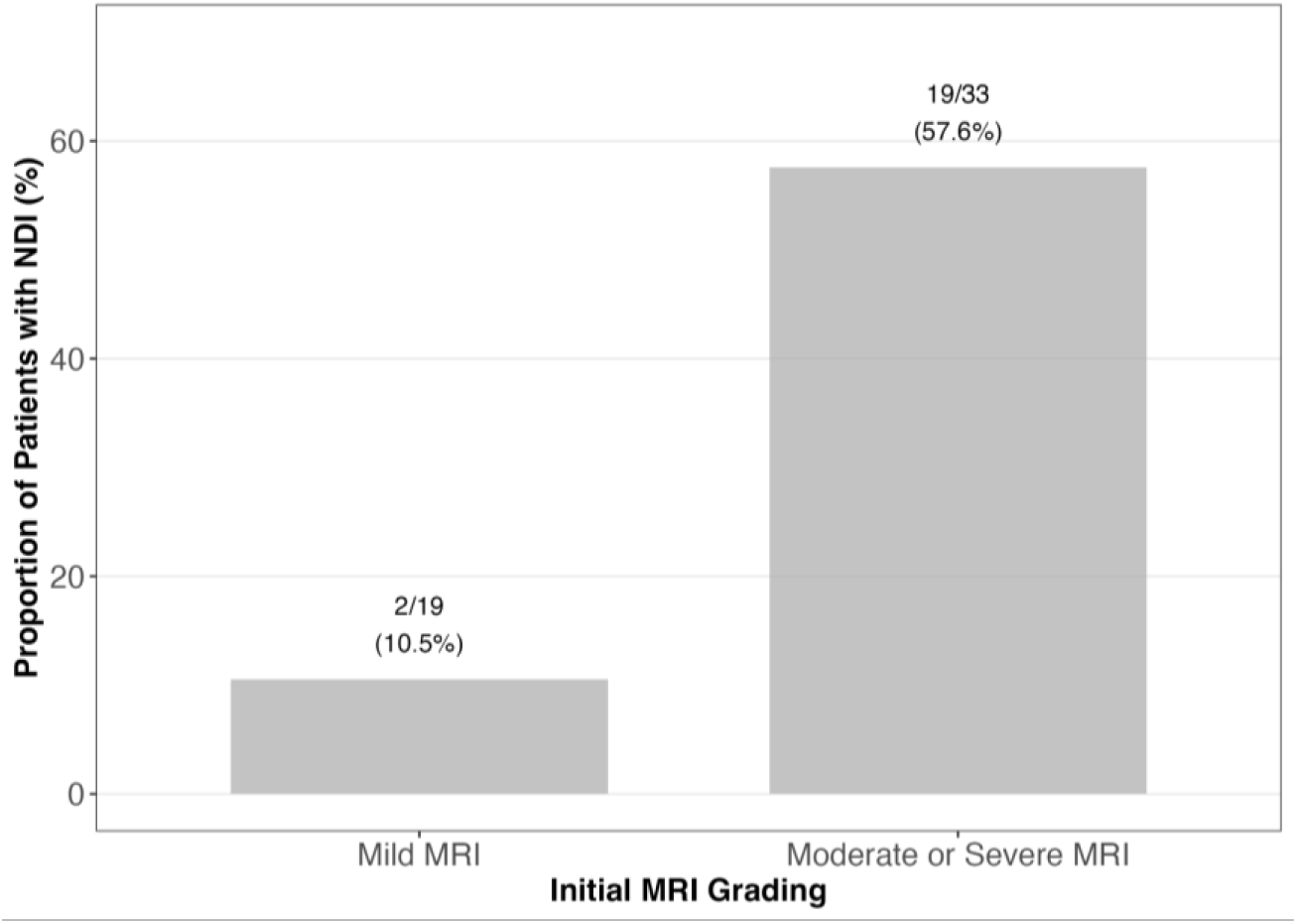
Proportion of NDI by MRI

**Figure 3:**
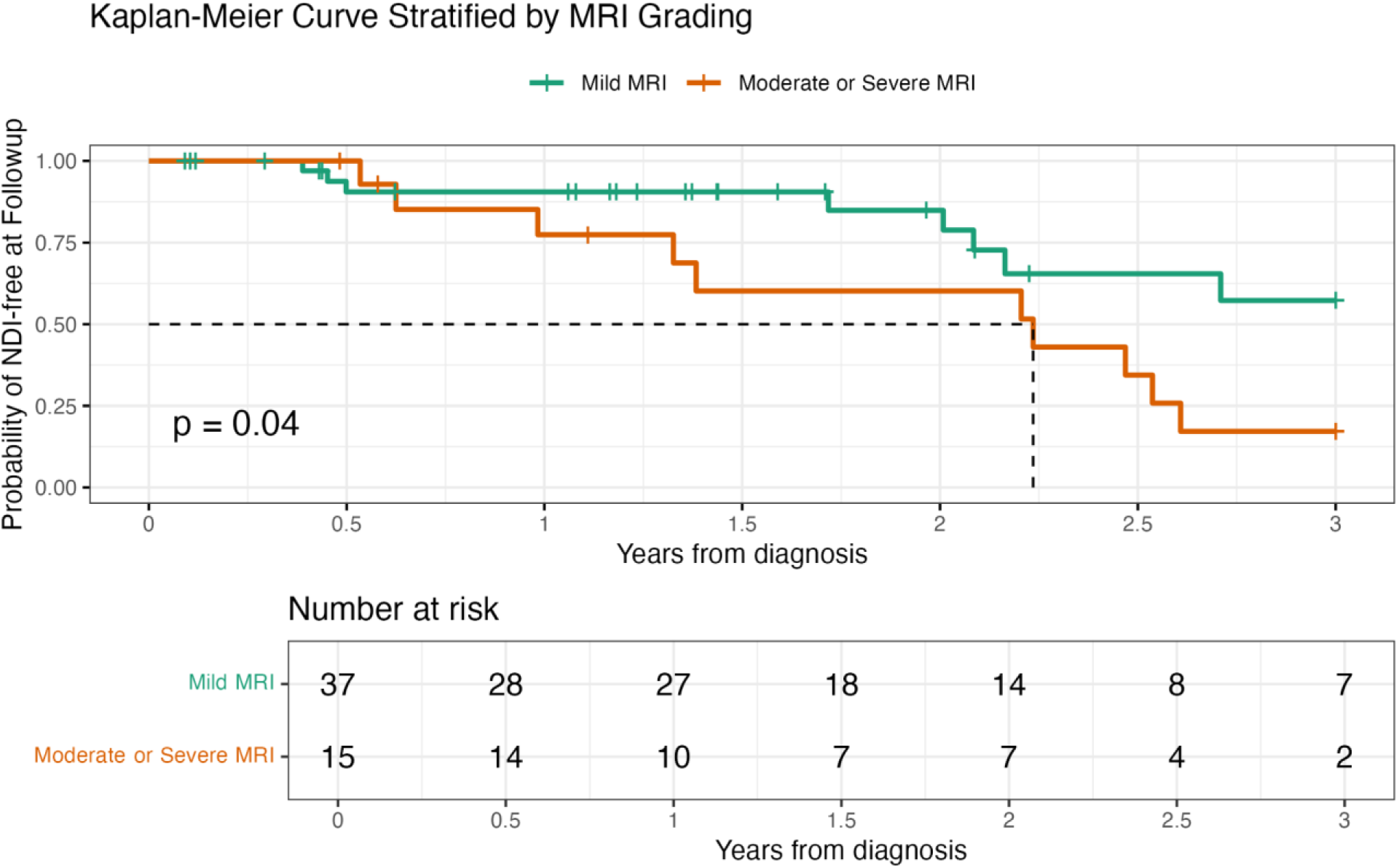
Kaplan-Meier Curve in Mild vs Moderate/Severe MRI

Our multivariate Cox proportional hazards model identified several independent risk factors of earlier development of NDI in our cohort (Table 4). Congenital heart disease emerged as the strongest risk factor, with affected infants exhibiting a 5.67-fold higher hazard of developing NDI sooner compared to unaffected infants (95% CI: 1.65 - 19.48, p = 0.0059). The presence of seizures at presentation was associated with a 4.32-fold higher risk of developing NDI sooner (95% CI: 1.32 - 14.17, p = 0.016). Infants with a 1-minute Apgar score ≥3 had a 3.12-fold higher risk of developing NDI sooner compared to those with scores <3 (95% CI: 1.062 - 9.14, p = 0.038).

**Table 4:** Multivariate analysis of potential risk factors of NDI (N=52)

| NDI Risk Factor | Odds Ratio (95% CI) | p value |
| --- | --- | --- |
| Initial MRI (Moderate or Severe vs. Mild) | 24.26 (4.70 - 180.16) | < **0.001** |
| Hypoxic-ischemic encephalopathy (Yes vs. No) | 5.81 (0.731- 61.54) | 0.11 |
| Congenital heart disease (Yes vs. No) | 4.98 (0.76 - 36.31) | 0.095 |
| Gestational age (in weeks) | 0.87 (0.65 - 1.10) | 0.31 |
| 1-minute APGAR score ($\geq 3$ vs. $< 3$) | 2.93 (0.44 - 26.72) | 0.29 |
| Seizures at Presentation (Yes vs. No) | 4.21 (0.78 - 30.83) | 0.11 |
| Cesarean delivery (vs. vaginal) | 1.24 (0.27 - 5.90) | 0.78 |

### Radiographic evolution

Twenty-nine of 63 patients (46.0%) underwent at least two MRI scans, with a median interval of 5.0 days (IQR 4.0-15.0) between scans. Most (82.8%, n=24) showed no change in MRI grade between scans. 10.3% (n=3) improved, while 6.9% (n=2) worsened. The mean grade change was minimal at −0.03 (SD 0.42) overall, with only slight variations among groups (Table 5).

**Table 5:**
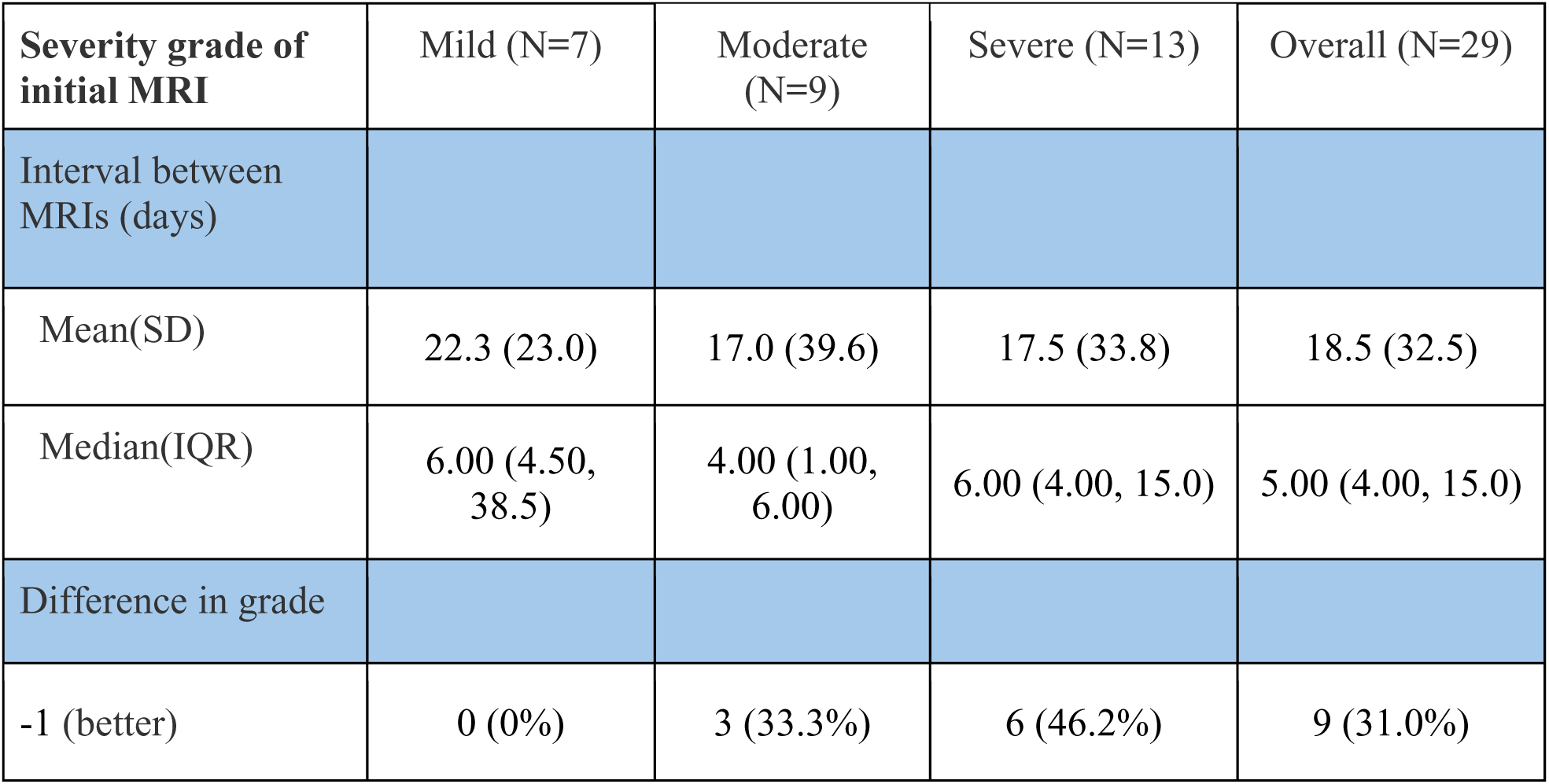
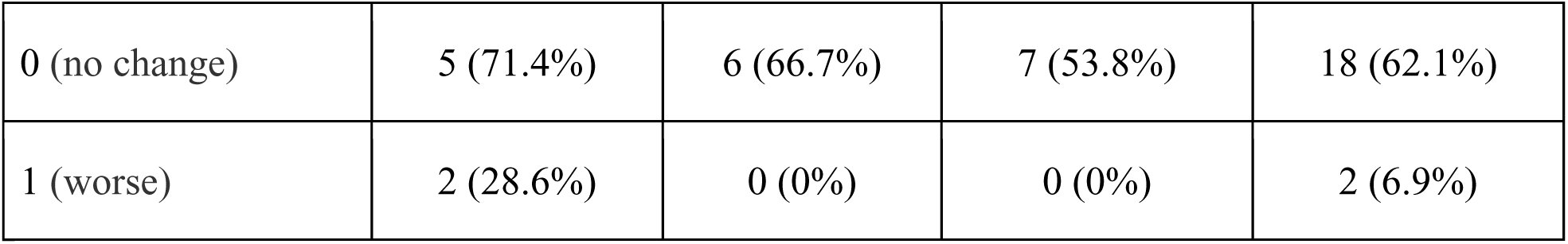
Interval grade changes between initial and first follow-up MRI (N=29)

| Severity grade of initial MRI | Mild (N=7) | Moderate (N=9) | Severe (N=13) | Overall (N=29) |
| --- | --- | --- | --- | --- |
| Interval between MRIs (days) |  |  |  |  |
| Mean(SD) | 22.3 (23.0) | 17.0 (39.6) | 17.5 (33.8) | 18.5 (32.5) |
| Median(IQR) | 6.00 (4.50, 38.5) | 4.00 (1.00, 6.00) | 6.00 (4.00, 15.0) | 5.00 (4.00, 15.0) |
| Difference in grade |  |  |  |  |
| -1 (better) | 0 (0%) | 3 (33.3%) | 6 (46.2%) | 9 (31.0%) |
| 0 (no change) | 5 (71.4%) | 6 (66.7%) | 7 (53.8%) | 18 (62.1%) |
| 1 (worse) | 2 (28.6%) | 0 (0%) | 0 (0%) | 2 (6.9%) |

## Discussion

Neonatal deep medullary vein thrombosis is an increasingly reported entity; however, its risk factors, management, and outcomes remain poorly understood. Here, we report neurodevelopmental outcomes for a large case series of 63 neonates with DMVT across a wide range of initial radiographic severity. In total, 21/52 neonates had NDI at follow-up. We also show that a simple, rapid radiographic grade is strongly associated with the likelihood of developing moderate to severe NDI at 2 years of age. On multivariate linear regression analysis, a moderate-to-severe grade MRI conferred a 24-fold higher likelihood of developing NDI than survivors with mild injury. Radiographic severity was also strongly associated with NDI on survival analyses. The association between radiographic finding and outcome has been noted in previous work with a more complicated grading scheme;^11^ Our study builds on this work by providing clinicians with a rapid, easily implemented grading system that can meaningfully inform conversations with families about an infant’s possible developmental outcome. Our multivariate Cox regression also identified important clinical factors, such as congenital heart disease, seizures on presentation, and poor Apgars as drivers of NDI risk, providing further prognostic nuance. The finding that fewer than 10% of infants with a mild initial MRI develop NDI is also clinically useful as milder DMVT is increasingly identified incidentally on imaging studies obtained for other indications.

Our infants were largely term or late preterm, and there was a non-significant preponderance of male infants. Among maternal and neonatal comorbidities that are likely risk factors for CSVT (which is better understood), we found that maternal and neonatal prothrombotic disorders were rare in our population; while systemic infection and congenital cardiac disease were common. The substantially higher presence of congenital cardiac disease is a notable difference between our population and other published series of DMVT^5,6,11^ and may reflect the high surgical volume for CHD at our institution. Infants largely presented within the first week of life, with symptoms including seizure, apnea/respiratory distress, and encephalopathy, overall at comparable rates to those reported in previous series and a recent meta-analysis.^5,6,11,13^

We found a notable difference between the Cox regression and multivariate logistic regression results regarding the effects of clinical variables on NDI risk. While seizures, congenital heart disease, and low Apgar score did not reach statistical significance in the logistic regression model, they emerged as significant predictors in the Cox regression analysis. The Cox model accounts for the timing of NDI events, which occurred between 4.66 to 32.59 months after initial presentation, with right-censoring at 3 years (censoring three patients whose events occurred after this time point). The significance in the Cox regression suggests that seizures and Apgar scores may be associated with not only if the outcome occurs, but also how quickly it develops. This finding highlights how traditional binary outcome analyses may mask these time-dependent relationships when evaluating early-life factors.

The natural history and optimal management of neonatal DMVT remain unknown. In our cohort, 29/63 infants were reimaged at a median of 5 days (IQR 4.0, 15.0). Of those who were reimaged, only 2 had radiographic worsening (both from mild to moderate). While our study was not designed to evaluate for association between treatment (e.g., hyperhydration, anticoagulation) and outcome, none of the 7 neonates who were anticoagulated for other indications had significant adverse effects from the anticoagulation. Further research is urgently needed to identify the risk of thrombus propagation in DMVT and the utility and risks of antithrombotic therapy in this population, particularly in light of the updated recommendations regarding anticoagulation for neonatal venous sinus thrombosis.^14^

Our study has a number of strengths. Our cohort represents the largest published series of neonatal DMVT to date, and we were able to extract follow up data for the majority of participants. The use of a composite NDI outcome that aligns with other neonatology studies contextualizes our findings within the broader field of neonatal brain injury. Our Cox regression analysis allowed us to identify additional time-dependent associations between infant comorbidities and developmental outcome, highlighting the dynamic, ongoing interactions between neonatal brain injury, comorbidities, and ongoing developmental processes. Finally, the inclusion of a sizeable number of infants with isolated DMVT or only punctate injury in the Mild Grade provides a broader view of how radiographic severity may inform outcome. This is relevant for practicing clinicians because isolated deep medullary vein congestion or thrombosis without associated parenchymal injury is increasingly reported on neonatal MRIs, and previous case reports or case series that have included only DMVT with associated parenchymal injury may not help inform discussions with the infant’s family about likely outcomes.

Because of its retrospective design, our study has inherent limitations. Not all survivors had follow up visits, although the distribution of radiographic grades was similar between those who did have follow up and those who did not. Because our primary endpoint (moderate to severe NDI at 2 years of age) was chosen to align with other research in neonatal brain injury,^12,15^ we were also unable to evaluate learning disabilities that might arise later in childhood. Similarly, this definition limits comparison of our findings to other studies of neonatal DMVT and CSVT that use different outcome measures, such as the Pediatric Stroke Outcome Measure (PSOM).^16^ A technical limitation is the inability to definitively differentiate isolated thrombosis from congestion, as both have similar radiographic appearance. It is also possible that results could be affected by the inclusion of both 1.5T and 3T scans, which was related to factors such as scanner availability and inclusion of scans from outside institutions in infants who were subsequently transferred to our hospital. Although our cohort had high rates of comorbidities that are common in neonatal CSVT, our study design did not permit us to identify whether these are risk factors for development of DMVT. Finally, due to study design and the small numbers of children who were treated with antithrombotic therapy, we were unable to comment on whether management choices such as hyperhydration or anticoagulation, which are used to treat neonatal CSVT, affected clinical or radiographic outcomes in our cohort.

Our study provides an easily implemented framework for clinicians to analyze acute imaging and counsel families about likely developmental outcomes for infants with DMVT. It also highlights important gaps that should direct future research in this area. In particular, a better understanding of the risk factors and acute management of DMVT are urgently needed.

## Data Availability

The data that support the findings of this study are available from the corresponding author upon request.

## Acknowledgements

The authors would like to thank Courtney Wusthoff, MD, MS for valuable insight regarding study design.

## Funding

The REDCap platform services at Stanford are subsidized by a) Stanford School of Medicine Research Office, and b) the National Center for Research Resources and the National Center for Advancing Translational Sciences, National Institutes of Health, through grant UL1 TR003142⤉.

## Disclosures

The authors do not have any relevant financial disclosures.

## Notes

### Competing Interest Statement

The authors have declared no competing interest.

